# Modeling of COVID-19 Pandemic and Scenarios for Containment

**DOI:** 10.1101/2020.03.27.20045849

**Authors:** Majid Einian, Hamid Reza Tabarraei

## Abstract

The impact of COVID-19 pandemic on the global health and world’s economy have been profound and unseen since the Spanish flu of 1918-19. As of now, many countries have been severely affected, partly because of slow responses to the crisis, ill-preparedness of their health system, and the fragile health infrastructure and the shortage of protective equipment. This note evaluates various scenarios, based on an estimation of number of identified and unidentified infected cases, and examines the effectiveness of different policy responses to contain this pandemic. Our result, based on an estimation of the model for Iran, show that in many instances the number of unidentified cases, including asymptomatic individuals, could be much bigger than the reported numbers. The results confirm that in such circumstances, social distancing alone cannot be an effective policy unless a large portion of the population confine themselves for an extended period of time, which is not only difficult to implement, but it could also prove extremely costly and damaging to the economy. An alternative policy, this note argues, is to couple effective social distancing with extensive testing, even to those who are asymptomatic, and isolate the identified cases actively. Otherwise, many lives will be lost, and the health system will collapse, adding to the ongoing economic troubles that many countries have started to encounter.

## 1 Introduction

As the impacts of the coronavirus—COVID-19—rattle the world on its path to more than 199 countries and territories (as of March 27), countries show astonishingly similar behavior, at least in the first phase, of this pandemic. Similar to some deadly diseases that affect individuals, it seems that the society and/or the governing authorities of many countries, go through the similar stages as individuals, namely denial, anger, blame, depression, and acceptance. We understand form scientists, that the sooner a patient accepts her sickness, the sooner she can receive treatment and care. A prolonged process to reach acceptance, makes it more difficult to effectively tackle the disease, and in many instances, if left untreated with unfortunate deadly outcomes.

For instance, Iran is a prime example of a country that has prolonged the denial phase, and in doing so, has created a situation that might lead to a catastrophic outcome and could result in a human tragedy. The authorities in Iran broke the news by announcing the death of two COVID-19 victims on February 19, 2020, just a day before nationwide parliamentary elections. Although it is almost impossible to know the exact date and the number of infected people prior to the official announcement, the number of new cases suddenly jumped from zero on February 18 to around 150 cases a week later.

Iran initially refused to quarantine the holy city of Qom, the source city of COVID-19 in Iran, and let the virus spread all over the country, and in a matter of a few weeks, all cities reported new cases of infected patients. Today, the pattern of pandemic in Iran indicates that, even after one month, the country is *still* grappling with the early stages of this terrible diseases and is unable to move on to the stage which it accepts the reality and takes extra-ordinary but necessary measures to confront the spread of the virus.

The data pattern in Iran shows a high death rate at the beginning of the pandemic, which then declined and reached the global average after a couple of weeks. Similarly, the pattern of identified cases has been very different across countries. For instance, while the crisis started sooner in Iran than many other countries, infected cases rose quickly in Europe and the US, and surpassed those of Iran. This is in despite of complete shutdown in many European countries and many states in the US. Similar to many other countries, this might have been due to the lack of resources for testing and identifying infected individuals. This is why this note tries to account for identified as well as unidentified cases and simulate a model-based number of infected population.

We use a modified Susceptible, Exposed, Infected, and Recovered (SEIR) model to simulate the spread of COVID-19. Although the model is fairly sophisticated, it is easily understood and it is estimated with available public data. The results for Iran show that active cases might be significantly higher than officially reported by 3 to 6 times in an optimistic scenario of shelter-in-place policy of 75 percent of the population. In a scenario where half of the individuals confine themselves, still around three million individuals get infected of which 50 thousand will die.

Therefore, the under-reporting of infected cases, although unintentional, not only undermines the integrity of responses, but ultimately puts more people at risk as they go unrecognized and untreated and continue to spread the virus with no end in sight.

The note examines multiple policy scenarios and their effectiveness. Although the model is estimated for Iran, many countries are in similar stages of the pandemic or soon will be in the same stage and therefore, based on the results, extreme measures need to be adopted to save lives and stop further spread of the virus. Social distancing alone cannot stop this disease except if more than 80 percent of the population confine themselves for quite an extended period of time. However, if the number of testing increases massively, and countries’ authorities successfully isolate infected individuals, the pandemic can be controlled before the summertime, even with a less degree of confinement.

While the paper simulates various scenarios of containment, it does not address the economic cost of each scenario. In line with recent findings, the note argues that a fast and aggressive multi-layer intervention is needed to have a substantial dent of the transmission. What the paper calls for is a clear and transparent strategy by governments to implement social distancing and aggressive testing. The testing, should be targeted to geographic locations that the virus is wide spread, to airports and cruise ships, to road travellers, and anyone who has been in areas that are hot spot or has been in any contact with someone in affected locations over the past two weeks. The strategy should not “just” focus on those that show symptoms but should include “all” who might have been exposed. In this exceptional environment, governments can use temporarily a combination of location data and credit card information to track COVID-19 in their countries and use surveillance tools to monitor quarantined individuals.

## 2 Model

### 2.1 The baseline model

Deterministic compartmental models are simple yet powerful models in describing this kind of epidemic outbreaks. We developed a variant of the SEIR model for the epidemic dynamic and control of COVID-19, using seven state variables and a minimal number of parameters to avoid over-fitting. The model is estimated for Iran, but it can be estimated for other country examples. It is different from a standard SEIR model by distinguishing hidden and identified infected cases. Indeed, standard SEIR models are estimated assuming that all infected people are reported. Such an assumption for the novel coronavirus pandemic is largely unreasonable, as many infected people show no or mild symptoms and as the testing procedure is not available in mass, many remain undetected. Thus, we divide infected compartment into Infected but not reported (I) and infected, detected and reported (Q) groups. We assume that identified cases are less infectious by taking self-precautionary measures and/or due to hospitalization. While unidentified cases can largely continue to infect the rest of the population. The model, therefore, distinguishes between the confirmed infected individuals, which are observable in data, and the total infected individuals. In reality, the number of confirmed cases is a function of medical tests and the preparedness of the healthcare system.

COVID-19 has an infection capability during the incubation (pre-symptomatic) period. In addition, based on the detection method, there exists a delay between the time that an individual is infected and the time this person is detected. State variables and parameters of the model in equations (2.1)-(2.7) capture these characteristics. Figure 1 shows the schematic diagrams illustrating these states (compartments).

**Figure 1:**
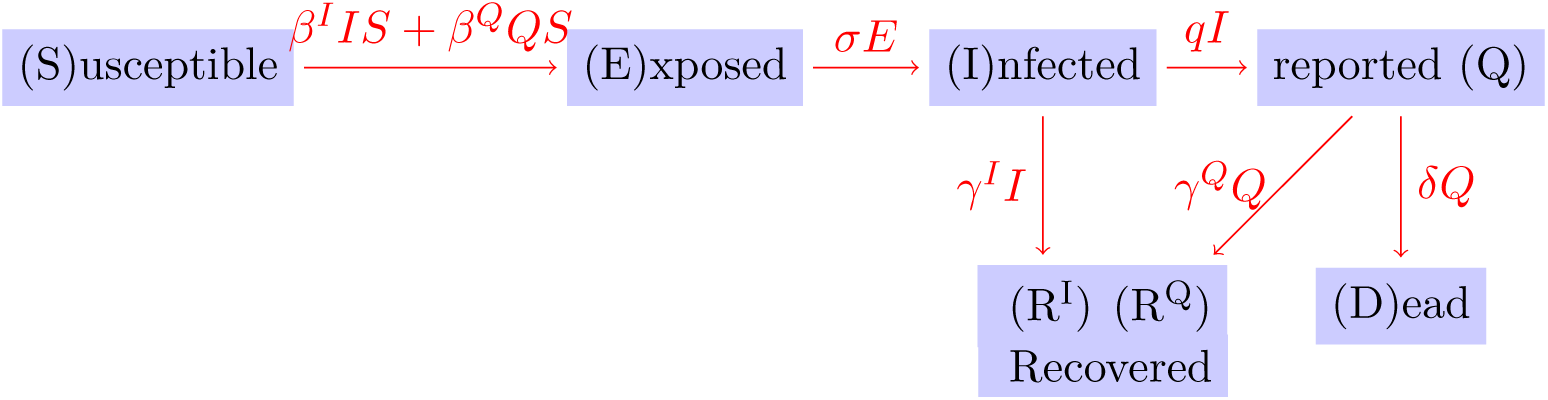
The schematic diagram of the phases of COVID-19.

We denote the share of susceptible individuals with no resistance to the disease in the population by *S*_*t*_. *E*_*t*_ is the share of people who are exposed to the virus but are not yet infected nor infectious. *I*_*t*_ is the infected population share who are undetected and hence un-quarantined. These individuals can be either symptomatic or pre-symptomatic. A share of these individuals, *q*, are detected and move to somewhat quarantined population *Q*_*t*_. Among both infected populations, 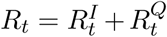 recover and for some time become resistant to new infections. Finally, we assume those who are detected are the individuals who need medical assistance and those that their general health condition might have deteriorated quickly. A share of these individuals, *δ*, pass away (*D*_*t*_).

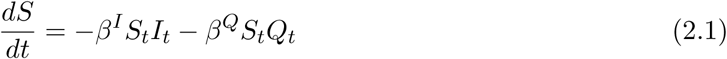

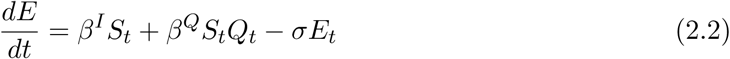

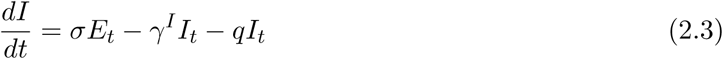

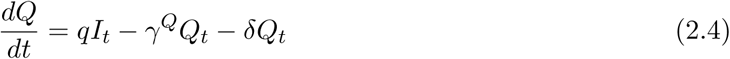

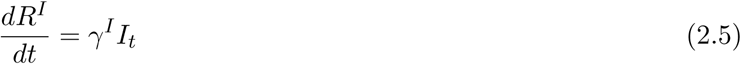

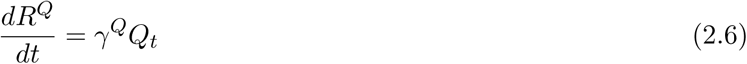

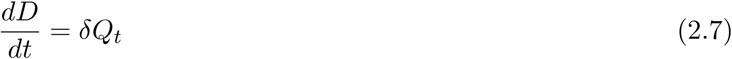

Finally, due to the high pace of the current outbreak, the model is a closed model in the sense that there are no new birth and death, except for the disease itself. In a simple SIER model, people are removed (hence the letter R in SEIR) from the analyzed population due to either immunity/resistance to the disease or death. Dividing the Recovered into “Recovered from *I*” and “Recovered from *Q*” groups, and keeping in mind that we only have data on recovered from *Q* and number of deaths (*D*), we can fit the model compartments to these series.

The values of *β*^*I*^ and *β*^*Q*^ depend on the contact rate between individuals and the probability of transmission in an encounter. 1*/σ* is the mean latent period and 1*/γ*^*I*^ and 1*/γ*^*Q*^ are the mean infectious periods for each group of patients. Finally, *γ*^*I*^ and *γ*^*Q*^ are the recovery rates and *δ* is the death rate among the *Q* group.

### 2.2 Estimation

the model is simulated by solving a deterministic Ordinary Differential Equations (ODE), given the parameters (*β*_1_, *β*_2_, *σ, γ*^*I*^, *q, γ*^*Q*^, *δ*) and the initial conditions (*S*_0_, *E*_0_, *I*_0_, *Q*_0_, 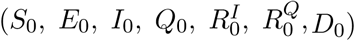. To obtain a realistic simulation of the baseline model and possible scenarios, we needed to estimate or calibrate these parameters and initial conditions. Since we are at the beginning of the epidemic cycle and the full distribution is not yet revealed, the model cannot be properly fit to a probability distribution function. As a result, the model is estimated by minimizing the differences between actual and predicted share of cases for observable data series which are *Q, R*^*Q*^, and *D* based on an extension of Drake (2011). The model is estimated using Iran’s official data for the number of confirmed, recovered and death cases^1^. As there are limited observable state variables, we fix or restrict a few parameters based on the reports in the literature to avoid over-fitting. First, based on the genomic studies on the phylogeny of SARS-CoV-2, two genetic mutations of the virus has happened in Iran (The Nextstrain team, 2020)^2^, first between January 10^th^ and January 27^th^, and the second between January 24^th^ and January 30^th^. Thus, the virus was present in Iran before January 27^th^, long before the first official record of the disease in Iran (February 19^th^), but not before January 1^st^ as the parent mutation has occurred in Hubei, China between January 1^st^ and January 14^th^. So, the starting time of the simulations should be between January 1^st^ and January 27^th^. Initial values for observable states are obviously zero. Second, we pin down the relationship between *β*^*I*^ and *γ*^*I*^, and between *β*^*Q*^ and *γ*^*Q*^, such that 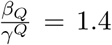, i.e., the minimum basic reproduction value and 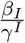 to be equal to maximum basic reproduction number of 6.6, based on various reported values^3^.

Third, the relationship between *γ*^*Q*^ and *δ* is fixed at 5.7 percent to match the case fatality rate with the one estimated by Baud et al. (2020). Without these restrictions, the parameters and initial states can be estimated, but the issue of over-fitting resides and leads to misspecification. Based on the optimization, the epidemic started on Jan 15^th^ with 1 infected and 290 exposed people.

## 3 Results

### 3.1 Baseline forecasts

In the absence of any control measures and/or changes in individual behaviors, we would expect a peak of infection (some of identified and unidentified patients) to occur around May 23, 2020 (figure 2). As discussed in the previous section, we fix 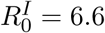 and 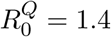 in this scenario for the whole epidemic period. Under this assumption, 70 percent of the population will be infected by around May 11^th^. As the epidemic progresses, at the peak, around 9 million identified individuals will be sick and will need medical assistance. In total, in an uncontrolled epidemic, around 900 thousand Iranians will die from this disease, not accounting for all other patients who will not get proper medical assistance during this period due to an overwhelmed medical system being.

**Figure 2:**
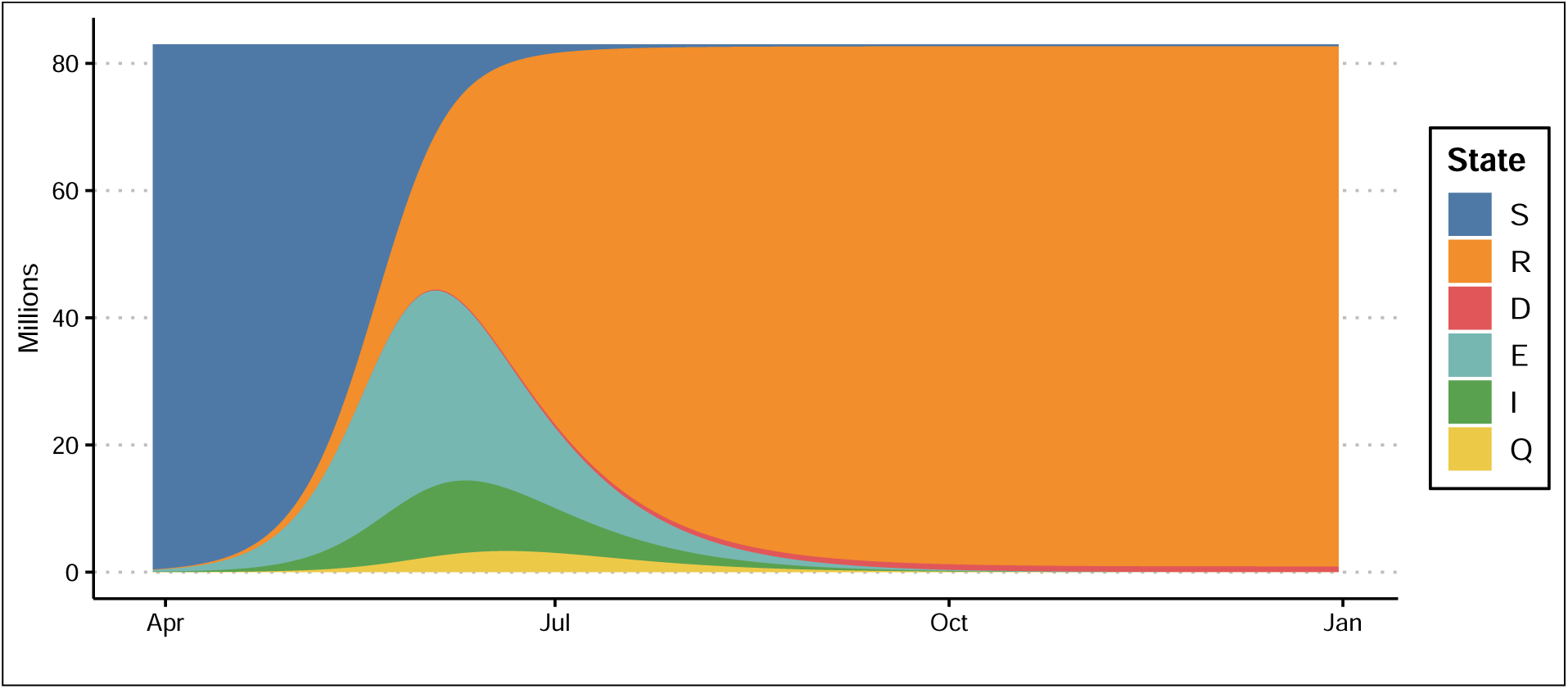
The baseline model forecasts.

In a cabinet meeting, Iran’s minister of Health mentioned that the maximum capacity of intensive care of all hospitals in Iran stands around 14,000 beds. Under the baseline scenario and without any control measures, at a 10 percent hospitalization rate of identified cases, all ICU beds in Iran will be filled by April 26 and even all hospital beds will be filled on May 26 (figure 3). Some 13,000 individuals will die per day around Mid-June (figure 4).

**Figure 3:**
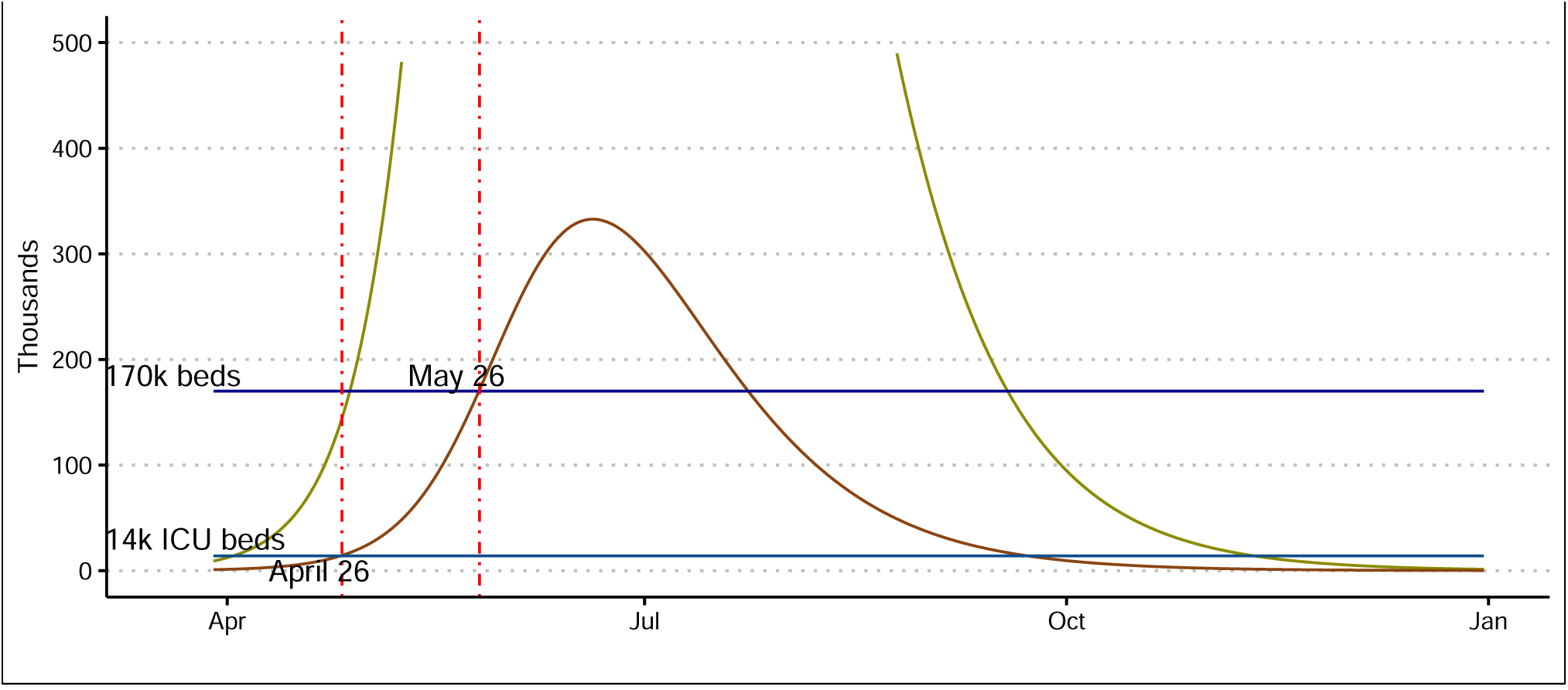
The baseline model forecasts: Identified cases needing hospitalization.

**Figure 4:**
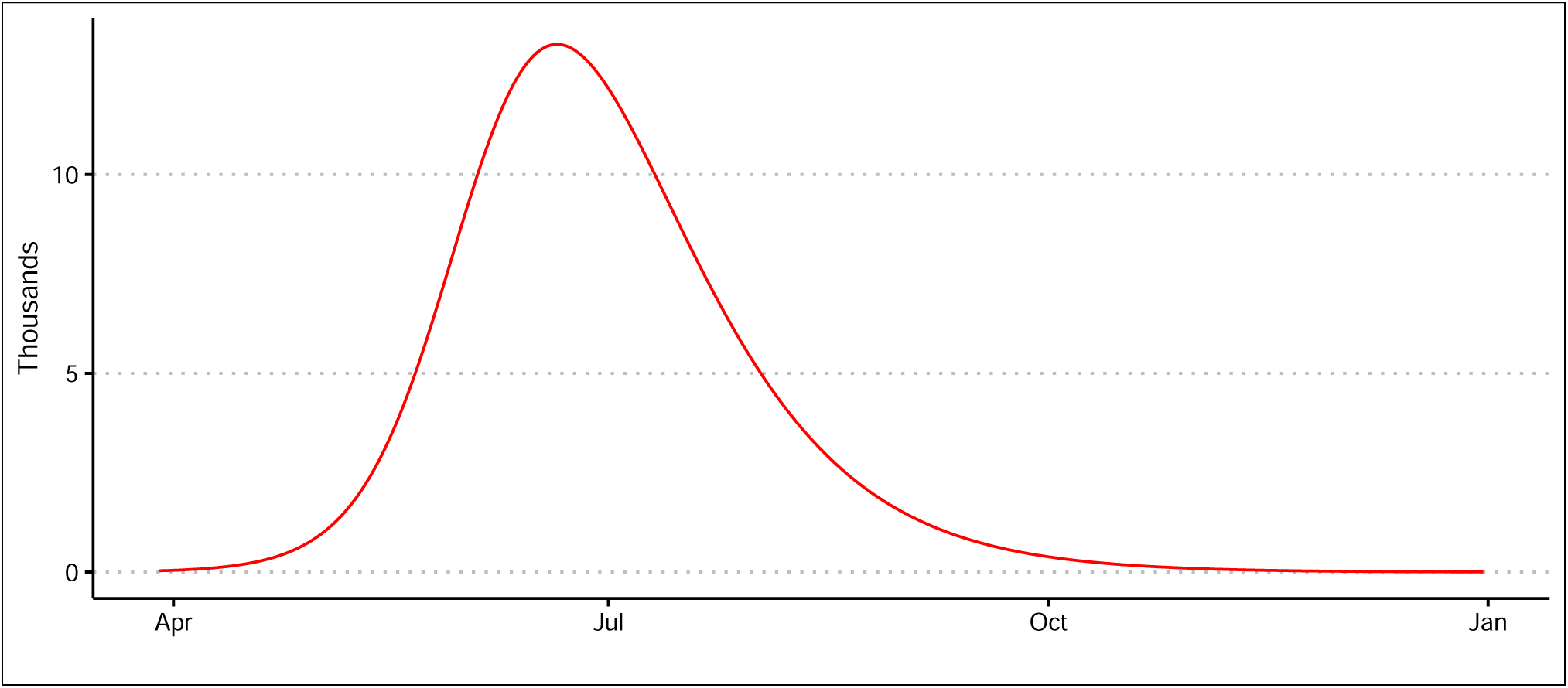
The baseline model forecasts: Deaths per day.

### 3.2 Scenarios

Although Iran initially refused to fully quarantine the holy city of Qom and later the other cities, the government started to react to the epidemic by cancelling all cultural, religious and sportive events, and closing of universities and schools in several cities. The authorities refused to quarantine areas affected by the outbreak and instead asked people to remain inside their houses on a voluntary basis, without any enforcing measures. Some religious sites remained open until mid-March. Unlike every year in late March (Persian New Year) the authorities urged the public to avoid travelling for the two-weeks holidays. However, according the official statistics, roads were still busy, and millions of people travelled, making the self-quarantine policy ineffective. In the absence of firm suppression policies^4^, we simulate the model based on the changes in the behaviors of the public by comparing *i*) the baseline or uncontrolled scenario, *ii*) with complete isolation of identified infected cases, *iii*) case isolation and self-quarantine, *iv*) intensive testing, identification of cases and quarantine, *v*) a combination of previous scenarios, *vi*) abandoning shelter-in-place policy one month after the peak of the epidemic under scenario *v*. Scenario (*ii*) is implemented by assuming *β*^*Q*^ *=0*, meaning that as soon as an individual is reported as infected, all necessary measures are taken to stop contagion from this person to other people. China forced hospitalization of all “confirmed cases” and not just those who required hospital care, to reduce onward transmission in the household and potentially in other social settings. Results show that this scenario is not effective and the epidemic progress almost at the same rate with and without complete isolation of identified cases.

Scenario (*iii*) is implemented by choosing smaller values for *β*^*I*^ of the base scenario, as it depends on the contact rate between individuals, in addition to *β*^*Q*^ *=0*. Adding social distancing saves many lives (figure 6), but at this stage of the epidemic at least 80 to 90 percent of the population should practice shelter-in-place. Sadly, if only one quarter of the population continue regular contacts with each other, the number of death cases can easily surpass a quarter of a million, and clearly, this policy will not be enough to overrun hospitals’ maximum capacity (figure 5).

**Figure 5:**
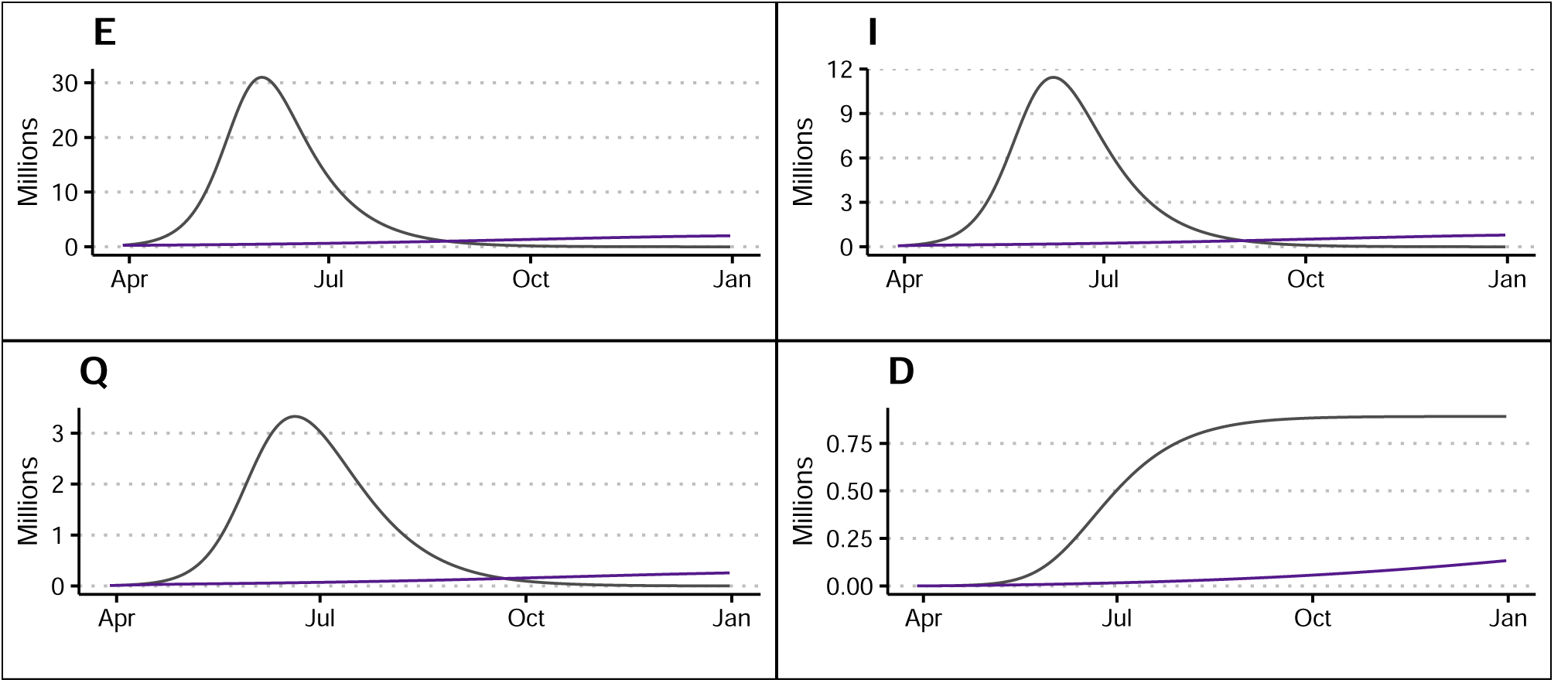
Baseline scenario (black) versus 75 percent shelter-in-place (purple).

**Figure 6:**
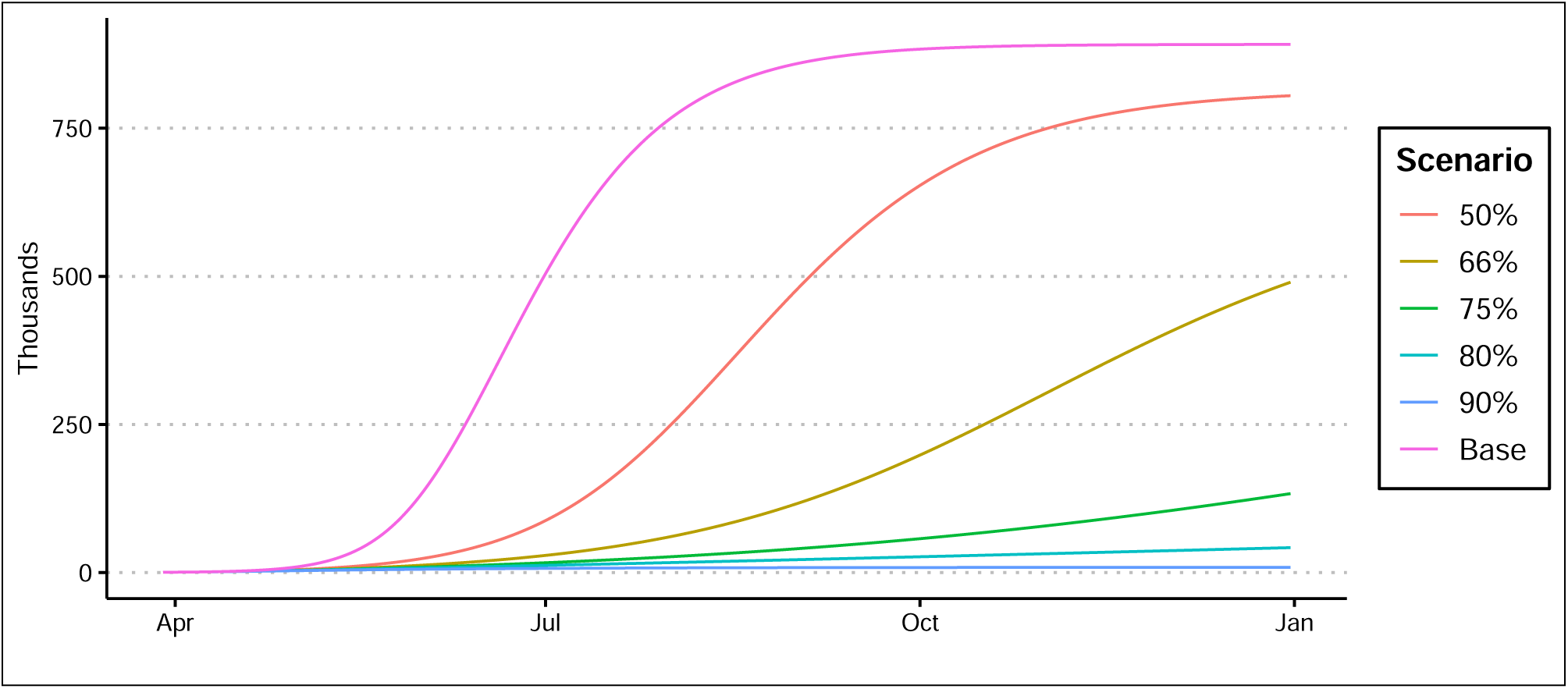
Total death count in case isolation and partial self-quarantine in different levels.

We implement scenario (*iv*) by assuming that the health authorities will increase number of testing by 10 folds. This helps isolate infected cases, provides patients with necessary medical assistance and decelerates spread of the disease to the rest of population. This policy would shift individuals from *I* group to *Q* group, enforcing better implementation of precautionary measures and control of contagion. The results are shown in figure 7. Even in this case, more than 75 percent of the population will contract the disease and the death toll will remain very heavy. Only by combining this scenario with previous scenarios, one can be hopeful to control the epidemic and save lives.

**Figure 7:**
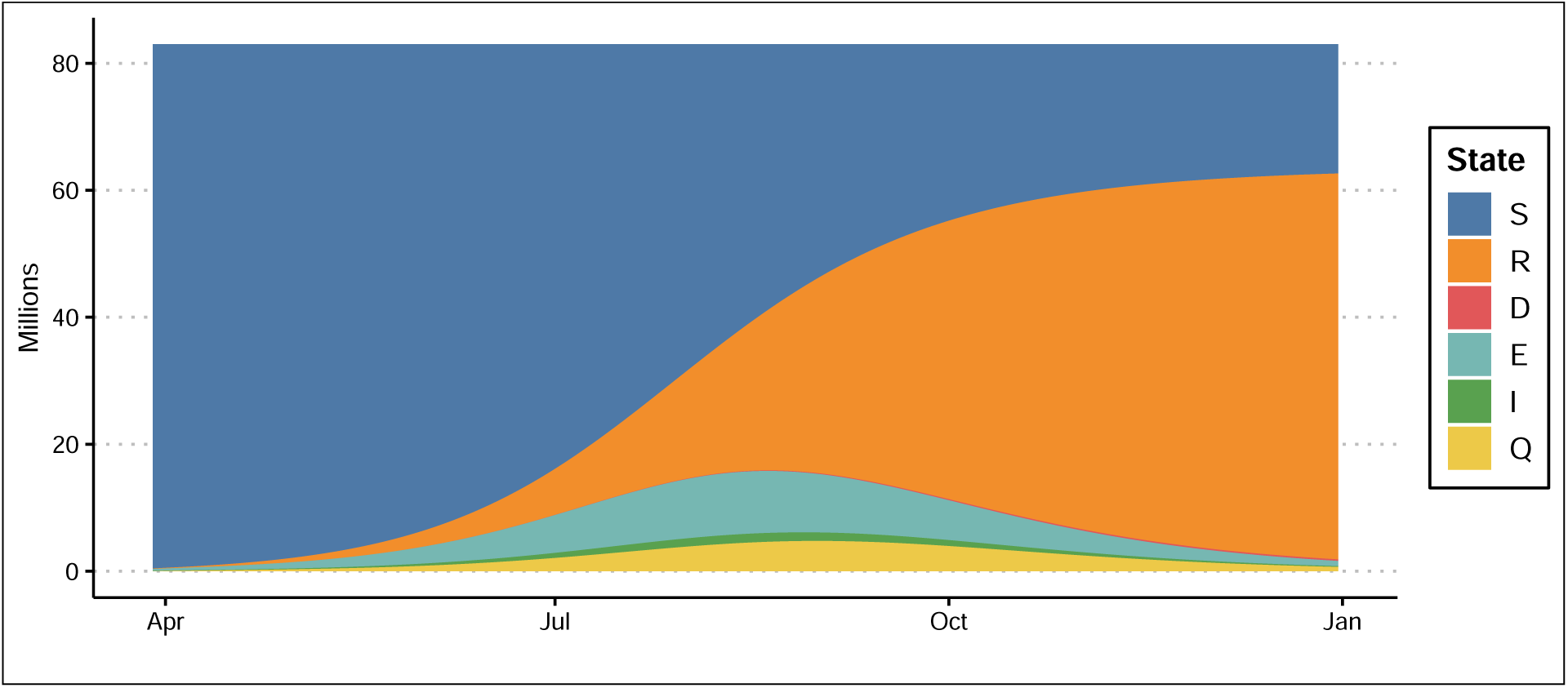
Increase testing by 10 times compared to the uncontrolled scenario, no case isolation and shelter-in-place

Scenario (*v*) shows the result for the combined policies, assuming that only 50 percent of the population respect the shelter-in-place policy, while testing and identification process intensify by 10 folds. Figure 8 shows the results for the combined policy. If implemented immediately, the total number of deaths by end-June will remain below 5 thousand individuals. The maximum number of infected cases (*I + Q*) will reach its maximum at 175,000 on May 9^th^ and the majority of the population will not contract the virus.

**Figure 8:**
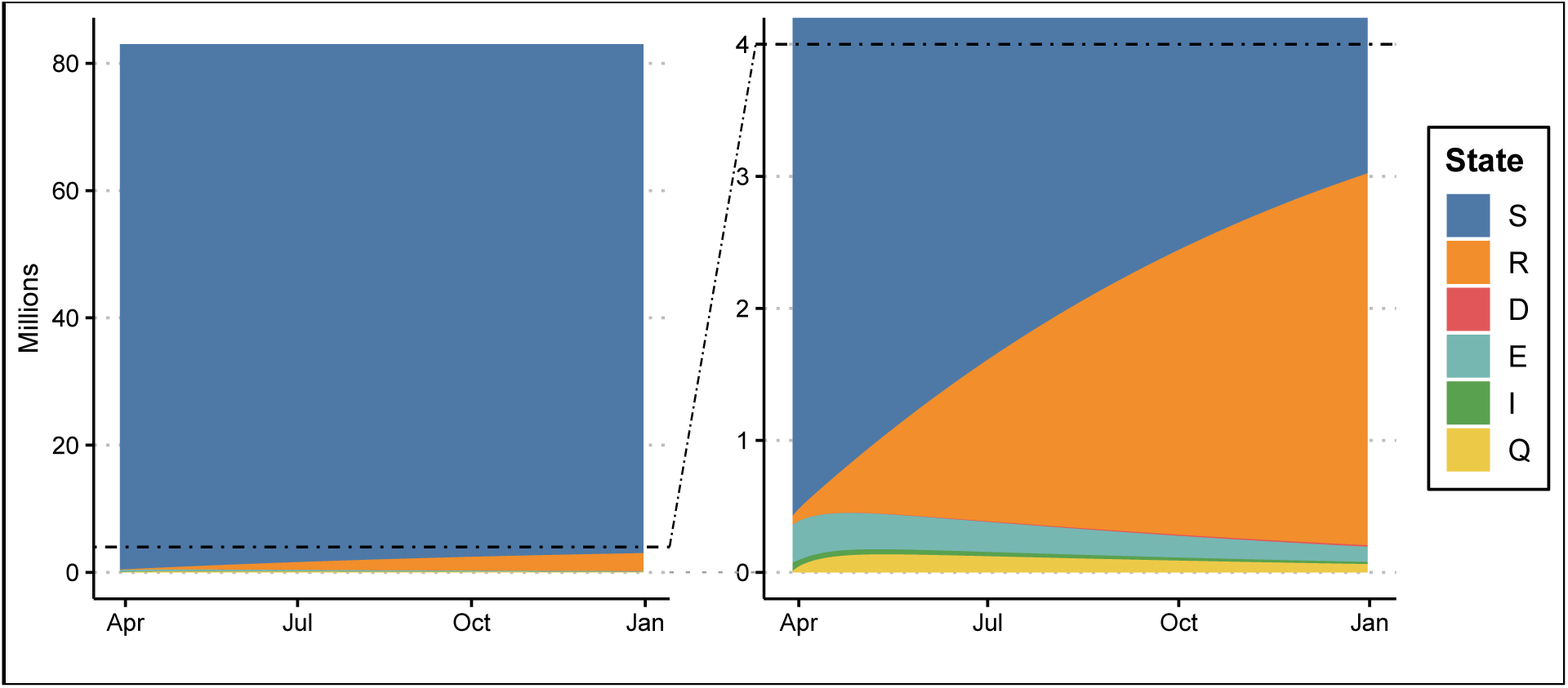
Combined scenarios: Intensive testing and shelter-in-place of 50 percent of the population

**Figure 9:**
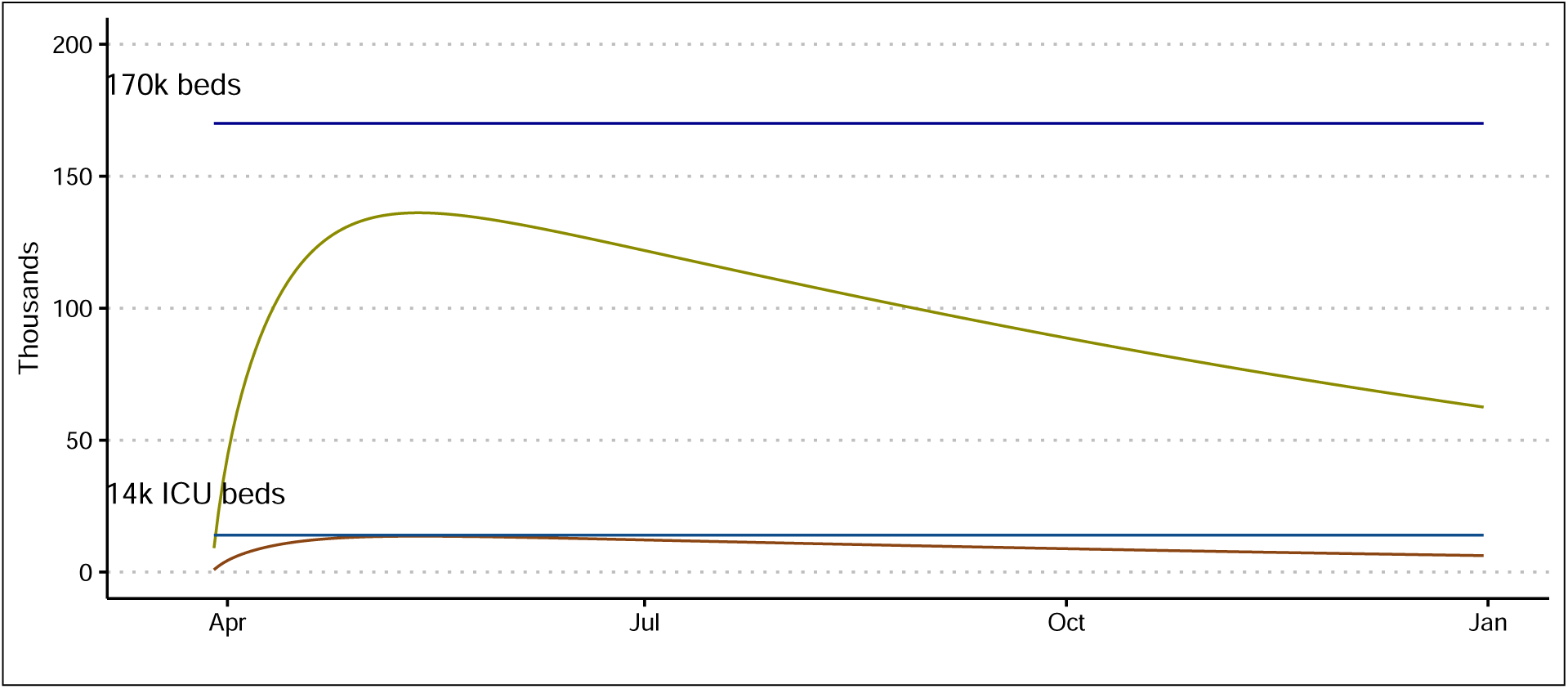
Combined scenarios: identified cases. The brown line shows the number of patients with intensive care needs (at 10 percent of total identified cases). Under this scenario, the peak number of patients remains below the hospitals’ ICU beds capacity.

**Figure 10:**
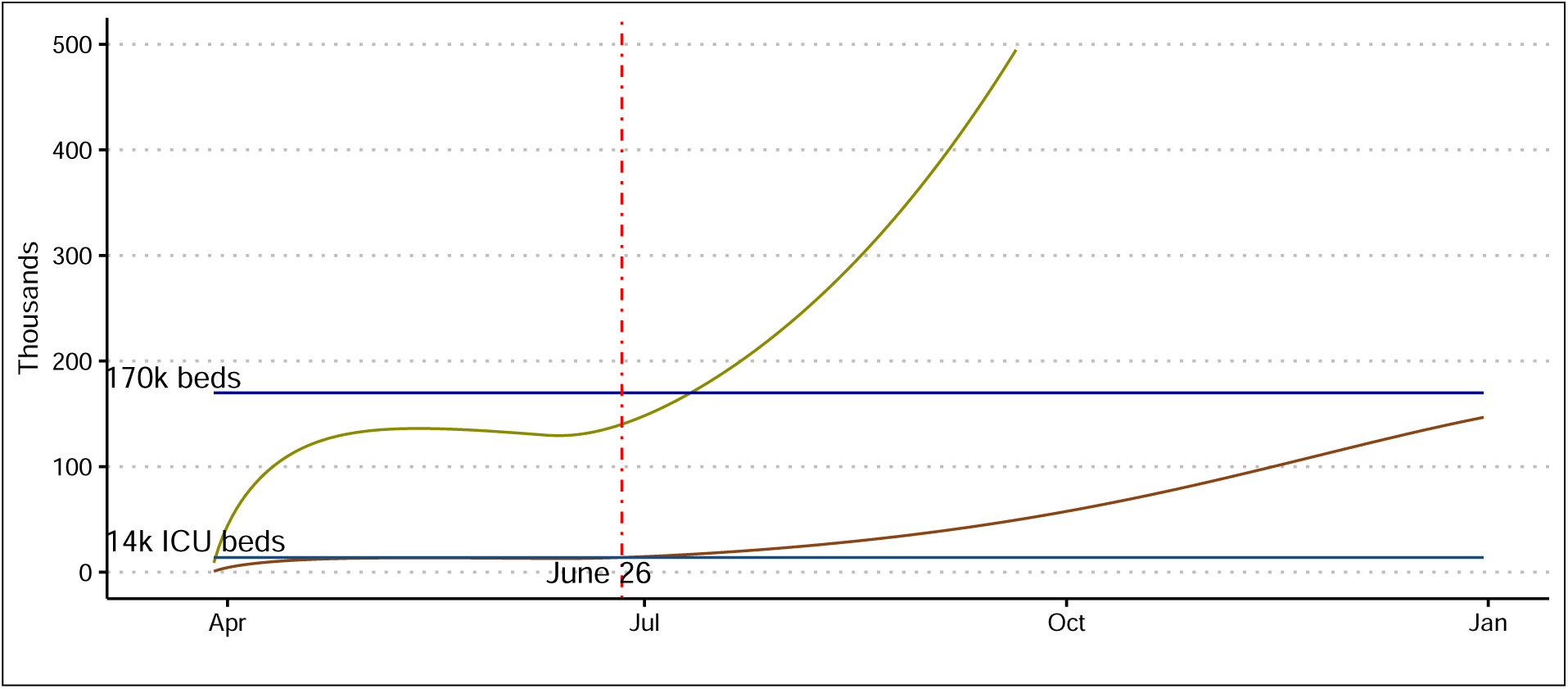
Ending the shelter-in-place policy 30-days after the peak of scenario (*v*), assuming that intensive testing continues and social interaction diminishes by 25 percent.

Finally, in the last scenario, we assume that last set of policies were effective enough to reach the peak of the epidemic on May 9 and the shelter-in-place policy continues by an additional 30-days period. As a result, businesses and government will open on June 9, while the intensive testing continues. We assume that as the shelter-in-place policy ends, all individuals remain vigilant and take hygiene measures. Under this scenario, social interaction reduces by 25 percent compared to normal times. As a result, the number of patients in need of intensive care surpasses the total ICU beds capacity before the end of June, while it remains below the total number of hospital beds in the country until the end of year.

## 4 Discussion and Policy Recommendations

Table 1 reports the number of death and recoveries at end-2020. Obviously, the combined set of policies, described in scenario *v* yields the best outcome with the lowest fatality and infected cases. Comparing scenario *iii* and *iv*, we conclude that a combined intensive testing, identifying infected individuals, and quarantine is a more effective policy than just a shelter-in-place policy. As discussed, scenario *vi* is the same as scenario *v*, assuming that the authorities end the shelter-in-place policy 30-days after the peak of the pandemic. Unfortunately, under such scenario the death toll can be still multiplied by 4 times compared to the scenario where social interactions are just reduced and kept low by 50 percent. If the social interaction is increased to normal times, the death toll can be 15 times larger than in the scenario *v*.

**Table 1:**
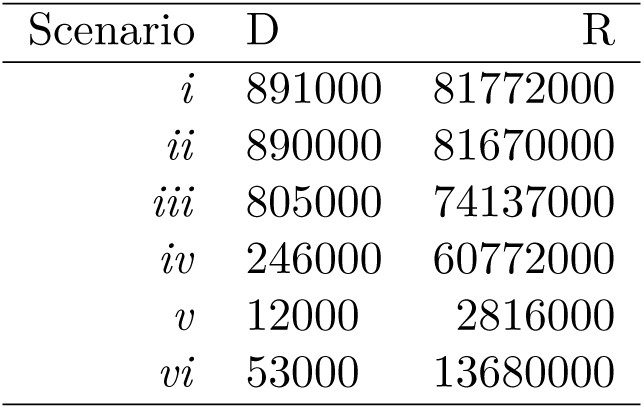
End of 2020 results

Many governments are impatient to open their countries businesses. They are rightly worried about the economic cost of a prolonged shutdown of economic activity. There are already outcry across countries by those who have lost their jobs. Many in low income and developing countries cannot afford to stay in home for long, since there are not enough safety nets to protect the most vulnerable. On the other hand, even in advanced economies, the ripple effect of a lengthy and uncertain shutdown of the economy is too painful. This is the reason that countries’ authorities have to start implementing a broad range of decisive actions.

Such policies should be put in place to save lives of thousands of people, alleviate pressures on the health system, and provide an opportunity for the people to go back to a normal living setting. These comprehensive set of measures include isolation of infected cases, shelter-in-place, and intensive and extensive testing. Good examples are Germany, South Korea and not surprisingly China, where it not only hospitalized all cases, but also enforced a population-wide social distancing to curb the contagion.

Unlike the current approach in some countries, where only the symptomatic cases are tested, the authorities should provide intensive and extensive testing and try to identify almost-all infected cases. This process would identify asymptomatic but infected cases and stop them of spreading the virus. This is key for a successful containment policy. Given the geographic distribution of the pandemic, the testing should focus on the most affected locations first and then expand to those people who have visited the hot spots or have been in contact with anyone in those areas, while everyone practice social distancing. A combination policy, as discussed above, not only reduces the number of infected, and ultimately the death toll, but it shorten the shelter-in-place period and consequently lowers the economic cost.

## Data Availability

All date used in the manuscript are public date, and the link to public data is provided in the footnotes of the manuscript. R codes used in the analysis will be provided by the corresponding author by request.

## Footnotes

1 Data source: 2019 Novel Coronavirus COVID-19 (2019-nCoV) Data Repository by Johns Hopkins CSSE https://github.com/CSSEGISandData/COVID-19

2 https://nextstrain.org/ncov?branchLabel=aa{&}label=clade:A3

3 Different reported values for the basic reproduction number for COVID-19 fall between 1.4 to 6.6. See Eisenberg (2020); Wu et al. (2020); Read et al. (2020); Liu et al. (2020); Riou and Althaus (2020); Li et al. (2020); Sanche et al. (2020)

4 Ferguson et al. (2020) defines suppression as a set of policies that aim to reduce the reproduction number to below 1

